# The effects of antibiotic prescribing for respiratory tract infection on future consultations in primary care: a systematic review and meta-analysis

**DOI:** 10.1101/2025.01.15.25320600

**Authors:** Ibrahim Adamu, Amanda Lambert, Safiyya Bello, Fatima Aminu Abdulmalik, Tom Marshall

**Affiliations:** Institute of Applied Health Research, University of Birmingham, B15 2TT; Professor of Public Health and Primary Care, Institute of Applied Health Research, University of Birmingham, B15 2TT

**Keywords:** Primary care, Antibiotics, Respiratory tract infection

## Abstract

**Objectives:** Prescribing antibiotics may reinforce patients’ beliefs antibiotics are needed and increase future consultations for similar symptoms. This review determines the effect of antibiotic prescribing for respiratory infections in primary care on future reattendance.

**Design, setting and participants:** A systematic review and meta-analysis of randomised controlled trials and cohort studies, following Preferred Reporting Items for Systematic Reviews and Meta-Analysis (PRISMA) guidelines.

Participants were adults or children presenting with respiratory infection in primary care.

**Interventions:** Prescription of antibiotics.

**Primary outcome measure:** Reattendance at least 28 days after initial consultation.

**Method:** Eligible studies included open-label randomised controlled trials (RCT) or cohort studies of antibiotics compared to no antibiotics in adults or children with respiratory infections. MEDLINE (Ovid), PubMed, Embase, the Cochrane Central Register of Controlled Trials, clinical trial registries and grey literature sources were searched from inception until 6^th^ February 2024. Two reviewers independently screened, selected, assessed quality and extracted data. Separate meta-analyses were presented for RCT and cohort studies.

**Results:** We identified 2128 records and reviewed 48 full texts, of which five met inclusion criteria. These reported three RCTs (1207 randomised to antibiotics, 672 controls) and three cohort studies (209,138 exposed to antibiotics, 46,469 controls). In the meta-analysis of randomised trials, relative risk (RR) of reattendance with antibiotics was 1.10 (95%CI: 0.99 to 1.23) and in the cohort studies RR was 1.21 (95%CI: 0.94 to 1.49). An important limitation is that most studies were in UK primary care.

**Conclusion:** Evidence suggests prescribing antibiotics for acute respiratory tract infections in primary care probably modestly increases future reattendance for similar conditions. Reducing antibiotic prescribing may help reduce demand for primary care.

**PROSPERO registration number:** CRD42023470731

**Article summary:** This systematic review of antibiotic prescribing for uncomplicated upper respiratory tract infections finds that antibiotics probably increase future consultations.

**Strengths and limitations:**

- This is the first systematic review of the effects of antibiotic prescribing for upper respiratory tract infections.
- The finding that antibiotic prescribing increases future consultations for upper respiratory tract infections is consistent with other evidence.
- The effect just fails to meet statistical significance.
- Almost all the studies were undertaken in the UK, which may limit the generalisability.

## Introduction

Although upper and lower respiratory symptoms are common in the UK, over 90% of those reporting sore throat, cough, cold or flu symptoms do not consult a general practitioner.^1,2^ Those who do consult are commonly prescribed antibiotics and consultations for respiratory symptoms account for most antibiotic prescribing in primary care.^3^ Indeed, changing rates of antibiotic prescribing are mainly attributable to changing rates of consultation (patient behaviour) and less by changing rates of prescribing for those who consult (clinician behaviour).^4^ Consultations for respiratory symptoms also contribute substantially to primary care workload.^5^

Patient belief in the efficacy of the doctor’s care and in the efficacy of self-care, are both strongly associated with the decision to consult.^6^ This suggests that when patients feel their complaint can be treated without consulting a doctor, they were less likely to consult. Patient expectations are also associated with antibiotic prescribing, although not as strongly as the doctor’s beliefs about those expectations.^7^ It may be possible for primary care practices to change patients’ beliefs and reduce consulting behaviour for respiratory conditions.^8^ Prescribing antibiotics for respiratory conditions may reinforce a patient’s belief that antibiotics are necessary for the condition, that they need to consult a doctor to obtain them, thus increasing future consultation rates for similar respiratory symptoms.^9^ If this medicalisation hypothesis is correct, prescribing antibiotics will increase future workload. The aim of this review is to assess the effects of prescribing antibiotics for respiratory infections in primary care on the frequency of future attendance for similar symptoms.

## Methods

A systematic review was conducted following the guidance of the Preferred Reporting Items for Systematic Reviews and Meta-Analysis.^10^

### Population, Intervention (Exposure), Comparison, Outcome and study types

Eligible studies included adults or children with respiratory tract infections in a primary care setting (the population of interest). Studies in secondary and tertiary care settings were excluded.

Respiratory tract infections include upper respiratory infections (the common cold, laryngitis, sore throat, pharyngitis or tonsillitis, acute rhinitis, acute rhinosinusitis and acute otitis media) and lower respiratory tract infections (cough, acute bronchitis, bronchiolitis, pneumonia and tracheitis).^11^ The intervention or exposure was prescription of any antibiotic (immediate or delayed, any dose or for any duration) and the comparator was no-antibiotic prescription. The outcome of interest was reattendance for respiratory tract symptoms at least four weeks after the initial consultation, this was to avoid including short-term reattendance for unresolved symptoms.

Eligible studies included randomised control trials, cohort studies and case-control studies with an open (unblinded) design. Patients must know if they received antibiotics for this to affect their beliefs in antibiotic effectiveness.

### Databases searched

The following databases were searched: MEDLINE OVID (1946 - 6^th^ February 2024), PubMed (1966 - 6^th^ February 2024), Embase (1974 - 6^th^ February 2024), Cochrane Central Register of Controlled Trials (CENTRAL) and Web of Science (inception to 6^th^ February 2024). (Supplementary Table 1, Supplementary Table 2, Supplementary Table 3, Supplementary Table 4, Supplementary Table 5).

Clinicaltrials.gov and clinicaltrialsregister.eu were searched for ongoing or recently completed trials and grey literature sources were searched (World Health Organisation, WHO website and the National Institute for Health and Care Excellence, NICE) were searched for relevant studies.

Reference lists of the included studies and relevant systematic reviews were checked for pertinent further studies.

### Screening and study selection

After the removal of duplicates, two independent reviewers (IA, SB, or FA) screened titles and abstracts against the eligibility criteria. Disagreements were resolved in consultation with a third reviewer. Full texts were retrieved of eligible studies and studies where eligibility could not be determined. Two independent reviewers (IA and TM) assessed the retrieved full texts against the inclusion and exclusion criteria. Two independent reviewers appraised quality of included studies using the Cochrane Risk of Bias 2 tool for randomised control trials,^12^ the Newcastle Ottawa scale (NOS) for cohort and case-control studies.^13^ The certainty of evidence was assessed using the GRADE criteria.^14^ Two independent reviewers extracted data from the included studies using a pre-specified data extraction form in Microsoft Excel. (Supplementary Table 6 and Supplementary Table 7) Records of excluded studies and reasons for exclusion were kept and presented. Disagreements on quality assessment or data extraction were resolved by discussion and with input from a third reviewer when consensus could not be reached.

### Meta-analysis

The primary meta-analysis was conducted for RCTs. A secondary meta-analysis including RCTs and cohort studies. Adjusted relative risks were used where available. Results were presented as forest plots. Stata 18 was used for analysis and production of forest plots. Heterogeneity was assessed using Chi^2^ and I^2^ statistics.^15^ Three pre-specified subgroup analyses were proposed: effect by study design (randomised controlled trials compared to cohort and case control studies); participant age (children compared to adults); the intervention type (immediate versus delayed antibiotics). If there were ten or more studies, publication bias was assessed using funnel plots.^16^ The review was registered on PROSPERO.^17^ Ethical approval was not sought as this is a review of secondary data sources.

### Deviations from protocol

The protocol specified a cut-off of two weeks to distinguish between short-term reattendance and long-term reattendance. After conducting initial searches, it was clear that most published studies used a cut-off of four weeks or 30 days. The definition of long-term reattendance was therefore modified to reflect this.

## Results

Database searches identified 2128 records after exclusion of duplicates. Of these, 38 met inclusion criteria and a further 10 records were identified from citation searches. Forty-eight full texts were reviewed, 43 were judged not to meet inclusion criteria, leaving five papers for inclusion. (Full texts included in Supplementary Table 8) The primary reasons for exclusions were: the outcome was short-term reattendance, relapse or non-resolution of symptoms (21); reattendance was not compared between groups receiving and not receiving antibiotics (9); reattendances were not reported (5); reported recurrent episodes of upper respiratory infections rather than reattendances (3); reported intention to consult (2); reported belief in the effectiveness of antibiotics (1); the intervention was an information leaflet (1); compared high and low-prescribing doctors (1). Some studies had more than one reason for exclusion. (See Figure 1).

**Figure 1:**
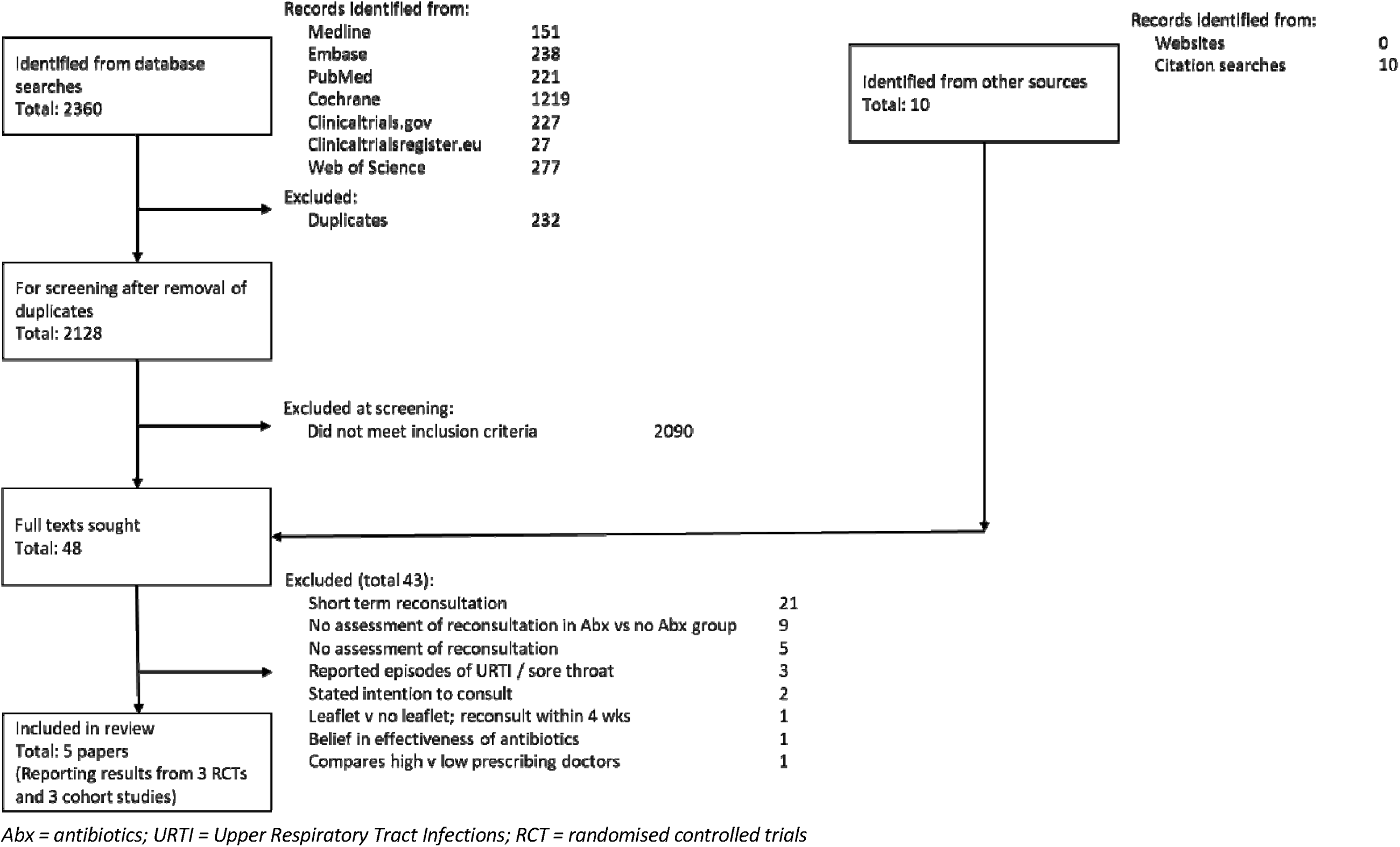
PRISMA diagram of included studies

**Figure 2.**
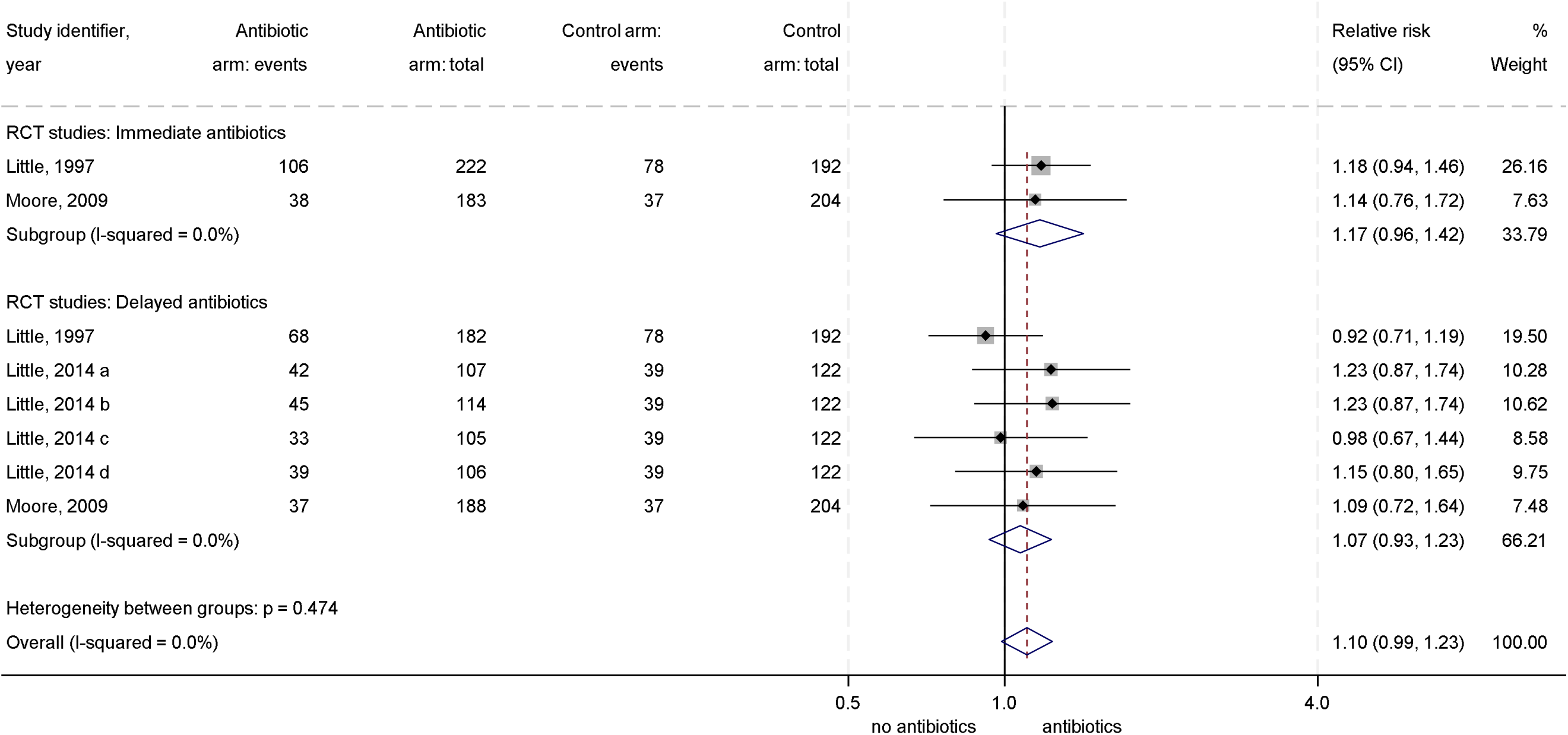
Forest plot of relative risk of reattendance for antibiotics versus no antibiotics, RCTs

**Figure 3.**
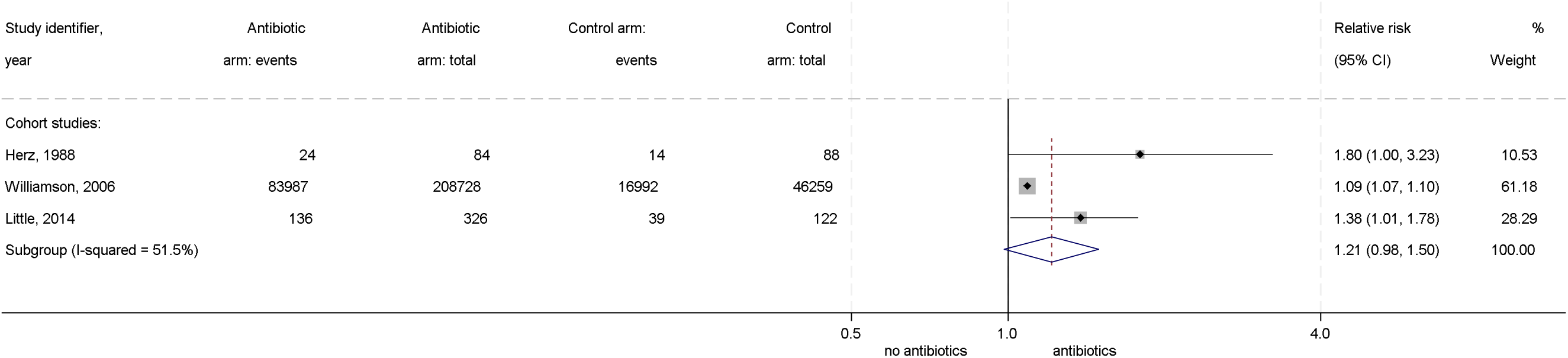
Forest plot of relative risk of reattendance for antibiotics versus no antibiotics, cohort studies

**Figure 4.**
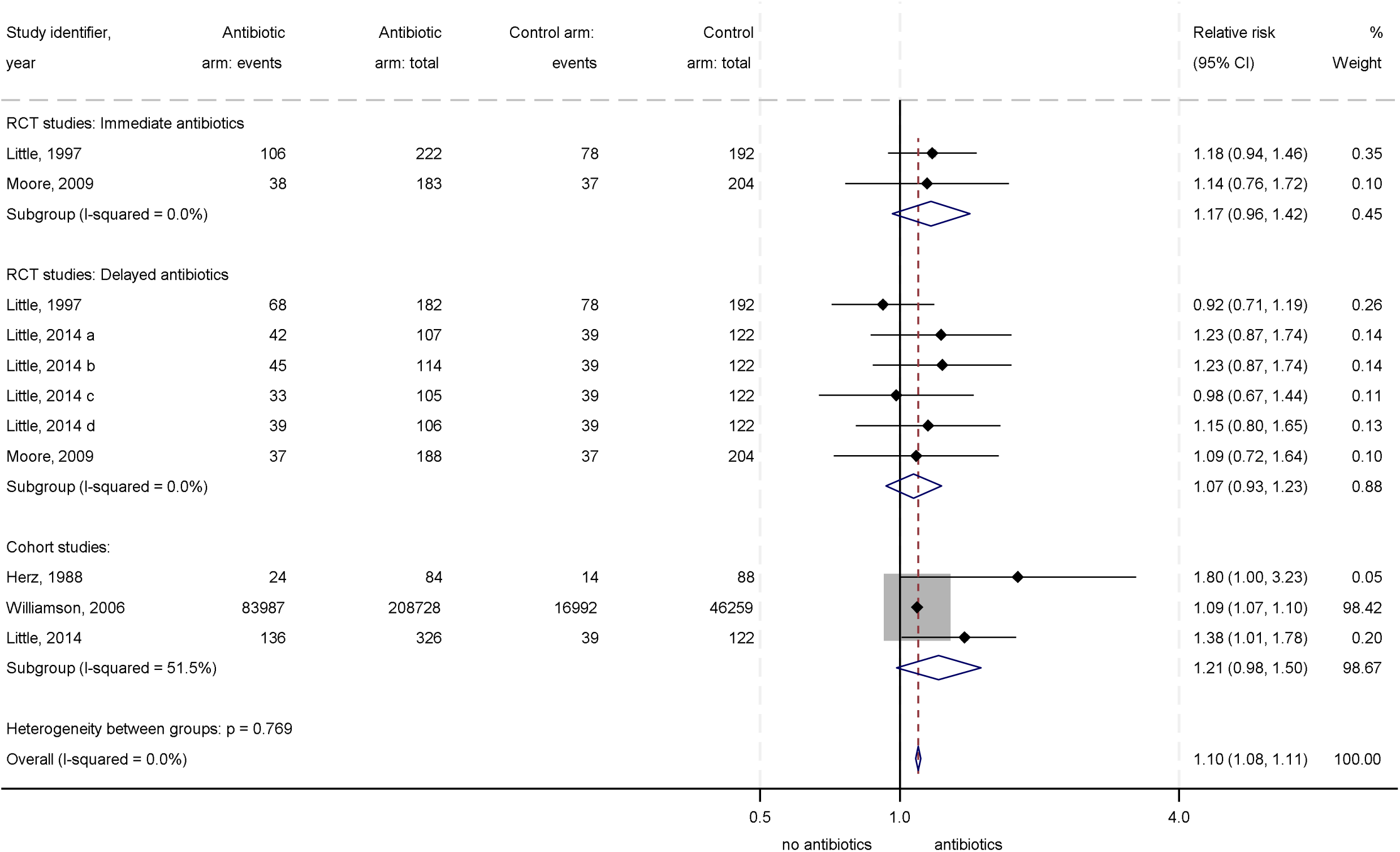
Forest plot of relative risk of reattendance for antibiotics versus no antibiotics, RCTs and cohort studies

The five papers reported findings from three RCTs and three cohort studies: one cohort study was reported in the same paper as a RCT. RCTs included 1207 participants randomised to either immediate or delayed antibiotics and 672 to no antibiotics.^9,18,19^ Three cohort studies included participants 209,138 exposed to antibiotics and 46,469 unexposed controls.^19,20,21^ (Table 1)

**Table 1:** Description of included studies.

| Study | Study design | Type of respiratory tract infection | Population | Country | Outcome | Intervention (antibiotic prescribing strategy) | Antibiotic arm: participants outcome / total | Control arm: participants outcome / total |
| --- | --- | --- | --- | --- | --- | --- | --- | --- |
| Little 1997 | RCT | Sore throat | Patients ≥4 years | England | Reattendance with sore throat 2 weeks to 1 year | Immediate prescription<br>Delayed prescription | 106/222<br>68/182 | 78/192<br>78/192 |
| Moore 2009 | RCT | LRTI | Patients ≥3 years | England | Reattendance with LRTI 1 to 12 months after the first visit | Immediate prescription<br>Delayed prescription | 38/183<br>37/188 | 37/204<br>37/204 |
| Little 2014 | RCT | Acute URTI and LRTI | Patients ≥3 years | UK | Reattendance with RTI after 1 month | (a) Delayed: phone practice for prescription<br>(b) Delayed: post-dated prescription<br>(c) Delayed: collect prescription from practice<br>(d) Delayed: patient decides whether to use prescription | 42/107<br>45/114<br>33/105<br>39/106 | 39/122<br>39/122<br>39/122<br>39/122 |
| Herz 1988 | Cohort study | Sore throat | Patients ≥21 | Israel | Reattendance after 1 month | Immediate prescription | 24/84 | 14/88 |
| Williamson 2006 | Cohort study | Acute otitis media | Any age | UK | Reattendance with acute otitis media at any time | Antibiotics prescribed | 83,987/208,728 | 16,992/46,259 |
| Little 2014 | Cohort study | Acute URTI and LRTI | Patients ≥3 years | UK | Reattendance with RTI after 1 month | Immediate prescription | 136/326 | 39/122 |

*RCT = randomised controlled trial; URTI = upper respiratory tract infection; LRTI = lower respiratory tract infection; RTI = respiratory tract infection*

### Randomised controlled trials

Little 1997 compared outcomes with no antibiotics, delayed antibiotics or immediate antibiotics for sore throat but did not report reattendance rates by antibiotic strategy. Data were obtained after contacting the authors. Moore 2009 compared no antibiotics, delayed antibiotics or immediate antibiotics for acute lower respiratory tract infections in a factorial design, which also evaluated the effects of providing a patient information leaflet. Little 2014 compared four delayed antibiotic strategies for acute respiratory tract infections: recontacting the practice by phone for a prescription, a post-dated prescription, allowing patients to collect the prescription themselves (collect) and prescribing but asking patients themselves to wait (patient led). The study also evaluated different strategies for symptom management in a factorial design and included a non-randomised comparison group of immediate antibiotics.

The randomised controlled trials were generally at low risk of bias. (Table 2) Selection of participants and randomisation processes were adequate; baseline characteristics were similar between allocated groups. There were relatively low rates of attrition and losses to follow up, outcome measurement and statistical analysis were appropriate. A change in the study protocol in Little 2014 to include an additional non-randomised arm was unlikely to have affected the findings. All three were conducted in UK primary care by the same group of UK researchers.

**Table 2:**
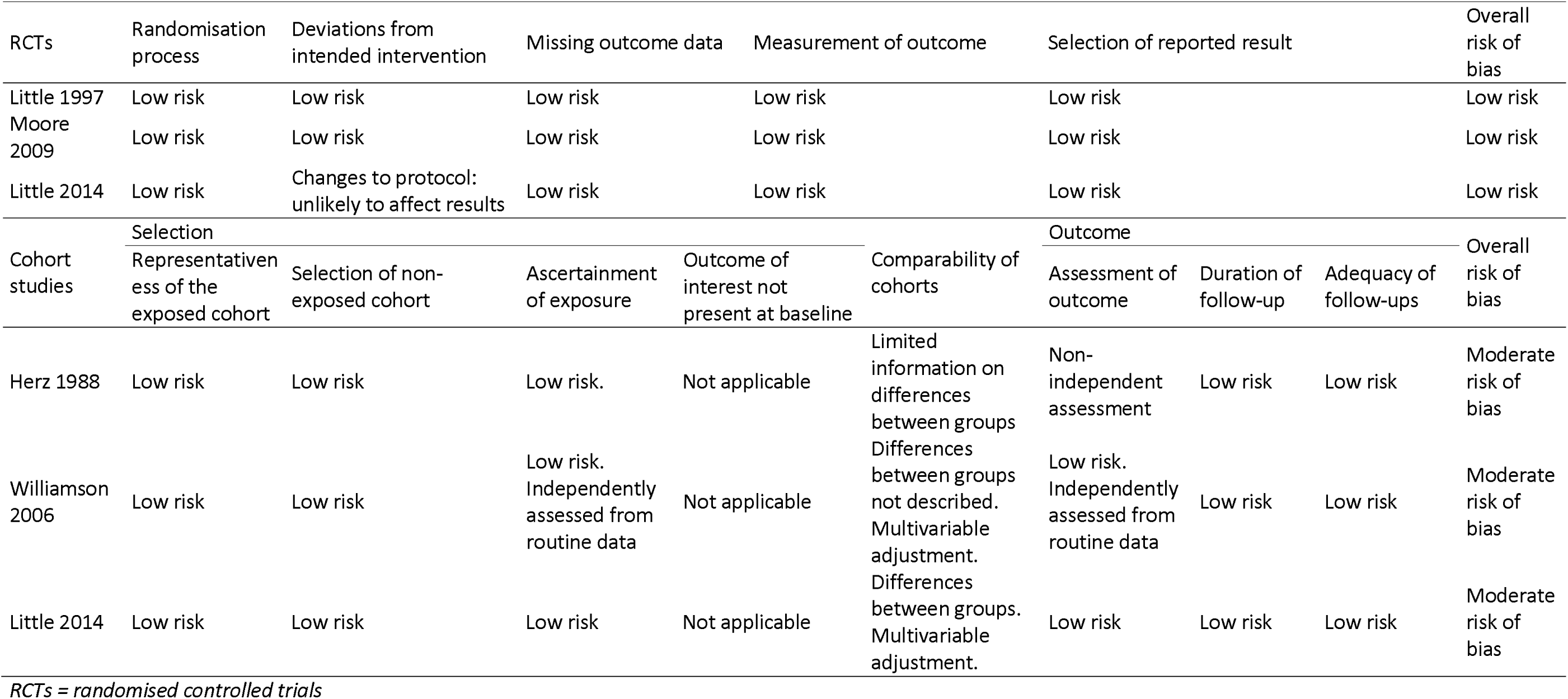
Risk of bias assessment of the included studies.

### Cohort studies

A retrospective cohort study in a large database of UK primary care records (Williamson 2006) compared reattendance rates in patients prescribed and not prescribed antibiotics for acute otitis media. Unadjusted hazard ratios and hazard ratios were adjusted for age, sex, multiple deprivation index, otorhinolaryngology referral and whether the patient was registered in high-prescribing general practice. Multivariable adjustment made only a small difference hazard ratio, suggesting those prescribed and not prescribed antibiotics were broadly similar.

One of the cohort studies was a non-randomised group reported in the same paper as a RCT (Little 2014). The non-randomised group was prescribed immediate antibiotics and compared to the group who received no antibiotics. At baseline these groups differed. The immediate antibiotics group had slightly more severe symptoms, was more likely to be labelled as lower respiratory tract infections and less likely to be labelled as having upper respiratory tract infections. Findings were adjusted for symptom severity, symptom control strategies, smoking, prior infections, gender, age and duration of follow-up.

In a small interventional cohort study, a primary care clinician in Israel prescribed antibiotics to 50 sequential patients presenting with sore throats, did not prescribe antibiotics to the next 102 patients and prescribed to a final 50 patients. (Herz 1988) The paper reported reattendances four weeks to six months after the index consultation. Neither prescribing nor reattendance were independently assessed and only limited information was provided on the comparability of the two groups.

The cohort studies were at moderate risk of bias, mainly because of differences between the groups at baseline. (Table 2) Two of the three cohort studies were undertaken in the UK by the same group of UK researchers as the RCTs.

### Meta-analysis

In random effects meta-analysis of the randomised controlled trials a relative risk (RR) of 1.10 (95% confidence interval (95%CI): 0.99 to 1.23) for reattendance was found in those randomised to antibiotics. There was no heterogeneity between included studies (I = 0%; Tau = 0). Three of the relative risks were calculated using the same control groups, thus were not independent. To test the effect of non-independence, a three-level random-intercepts meta-analysis of results within studies was carried out. This had no meaningful effect on the results (RR=1.09, 95%CI: 1.06 to 1.13) or on heterogeneity (I = 0%; Tau = 0). Random effects meta-analysis of both RCTs and cohort studies found a combined RR of 1.10 (95%CI: 1.08 to 1.11). (Figure 4). Random effects meta-analysis of cohort studies alone found a RR of reattendance in those receiving antibiotics of 1.21 (95%CI: 0.94 to 1.49), with moderate heterogeneity between the cohort studies (I = 51.5%, Tau = 0.02). (Figure 3).

Subgroup analysis by the age of patients was not possible as all but one study reported combined results for patients of all ages. Similar results were found for immediate (RR = 1.17, 95%CI: 0.92 to 1.42) and delayed (RR = 1.07, 95%CI: 0.93 to 1.23) antibiotic prescribing strategies compared to no antibiotics. (Figure 2).

Because acute otitis media might be considered clinically different from the other conditions, further sensitivity analysis tested the effect of omitting the large acute otitis media cohort study (Williamson, 2006). This had little effect on the findings when RCTs and cohort studies were combined (RR = 1.14, 95%CI: 1.03 to 1.27) (Supplementary Figure 1) (RR=1.15, 95%CI: 1.04 to 1.28 in the three-level meta-analysis) but increased the effect in the cohort studies subgroup to 1.39 (95%CI: 1.07 to 1.80) (Supplementary Figure 2).

## Discussion

### Summary

We found evidence that prescribing antibiotics for acute respiratory tract infections increases the frequency of reattendance for similar conditions.

### Strengths and limitations

Although RCT evidence alone did not reach statistical significance the findings were consistent between RCT and cohort studies, across various respiratory infections, for both immediate and delayed antibiotic prescribing strategies. Applying the GRADE criteria, we rated the certainty of RCT evidence as high because of the consistency of the findings.^14^ The generalisability of the findings is less certain as almost all of the included studies were conducted in UK primary care and all were conducted ten or more years ago. The cohort study evidence is at higher risk of bias than RCT evidence because of the possibility of unmeasured confounding, including confounding by indication, where antibiotics are prescribed to more severely affected patients. Nevertheless, both study types produced similar results.

### Comparison with existing literature

Other research has reached similar conclusions. A cohort analysis of 232,256 visits in 736 urgent care centres in the US compared repeat consultation rates in patients consulting high-prescribing clinicians, of whom 81% prescribed antibiotics, to low-prescribing clinicians, of whom 42% prescribed antibiotics. Those consulting high prescribing clinicians were more likely to reattend for acute respiratory infections: 68.8 versus 63.3 attendances per 100 (RR = 1.09, P <0.001).^23^ The authors noted that assignment to clinician was ‘quasi-random’. An analysis of 108 UK general practices over a six-year period found that general practices which reduce the proportion of acute respiratory infection consultations at which antibiotics are prescribed, saw subsequent reductions in acute respiratory infection consultation rates.^24^

### Implications for Research and Practice

The ways in which antibiotic prescribing affect future prescribing behaviour may differ for different types of upper respiratory tract infection. Prescribing antibiotics is hypothesised to affect patient belief in the effectiveness of antibiotics, intention to consult and therefore consulting behaviour. Several studies identified in the searches were excluded because they did not report consulting behaviour, but they found evidence of effects on belief and intention. Compared to no antibiotics, randomisation to immediate or delayed antibiotics increased the belief that antibiotics were effective for sore throat^25^ and for acute respiratory infections.^26^ A non-randomised comparison also found antibiotics increased belief in the effectiveness of antibiotics for acute respiratory infections.^19^ Random effects meta-analysis gave a combined relative risk of 1.19 (95%CI: 0.86 to 1.64) for belief in antibiotic effectiveness. In one study, an immediate prescription for antibiotics (where 99% received antibiotics) also increased the belief that antibiotics were effective for acute otitis media compared to a delayed prescription (where 24% received antibiotics).^27^ Three of these trials also reported patients’ intentions to consult in the future were higher in those prescribed antibiotics.^25,26,27^ Medicalisation is not the only mechanism by which antibiotics might increase future consultation rates. Antibiotics may also increase the risk of clinical recurrence. Long-term follow up of two double-blind randomised controlled trials of antibiotics in children, found those assigned to antibiotics may have higher rates of carer-reported episodes of acute otitis media (RR: 1.46, 95%CI: 1.08 to 1.97) and of sore throat (RR: 1.20, 95%CI: 0.80 to 1.78).^28,29^ Blinding eliminates the effect of carer beliefs about antibiotic effectiveness on carer-reported recurrences. Future research could investigate the contributions of beliefs and clinical recurrence to future consultation behaviour for different types of upper respiratory tract infections.

Despite the potential importance of prescribing as a cause of medicalisation, we found relatively few randomised controlled trials assessed longer-term changes in medicalisation or future consulting behaviour resulting from prescribing. This should be considered in all clinical trials of managing self-limiting illness in primary care. The predominance of evidence from UK primary care also points to a need for evidence from other settings. We did not address the question of whether immediate prescribing compared to delayed antibiotic prescribing has a similar effect on future consultations rates. This would require a search for studies with different intervention and comparison groups. The potential role of other types of discretionary prescribing on medicalisation and future consulting behaviour should also be explored.

### Implications for practice

It is plausible both that reducing antibiotic prescribing will reduce future consultations and that post-pandemic increases in antibiotic prescribing for upper respiratory infections may have increased demand.^30^ Practice policies which reduce antibiotic prescribing, such as dialogue with colleagues, consistent within-practice prescribing, supportive practice policies and increasing continuity of care may contribute to managing future demand for primary care.^31,32^

## Ethical approval

Ethical approval was not sought for this research as it is a secondary review of published research.

## Supporting information

Included papers

## Data Availability

Search strategies are included in supplementary materials.
The list of included papers is available from the authors on request.

## Acknowledgements

Thanks are due to Dr Beth Stuart and Professor Paul Little for providing original data from the Little 1997 study.

## Author statement

Ibrahim Adamu devised the search strategies, undertook searches and wrote a first draft of the paper. Amanda Lambert undertook statistical analysis and contributed to writing the paper.

Safiyya Bello contributed to the systematic review and to writing the paper.

Fatima Aminu Abdulmalik contributed to the systematic review and to writing the paper.

Tom Marshall had the original idea for the research, wrote the paper based on a first draft by IA.

## Data statement

Search strategies are included in supplementary materials.

The list of included papers is available from the authors on request.

---

**Supplementary Figure 1:**
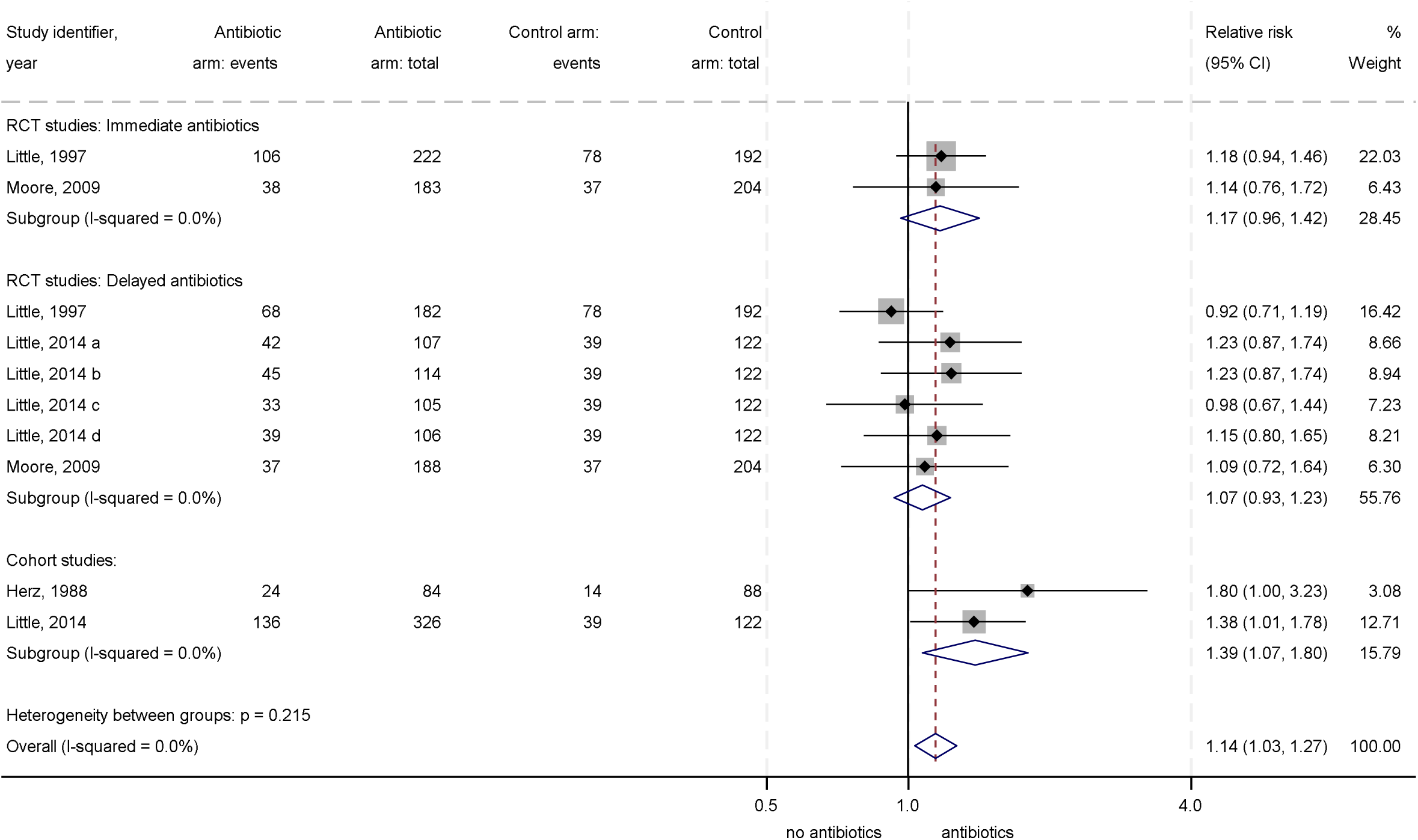
Forest plot of relative risk of reattendance for antibiotics versus no antibiotics, RCTs and cohort studies excluding Williamson, 2006

**Supplementary Figure 2:**
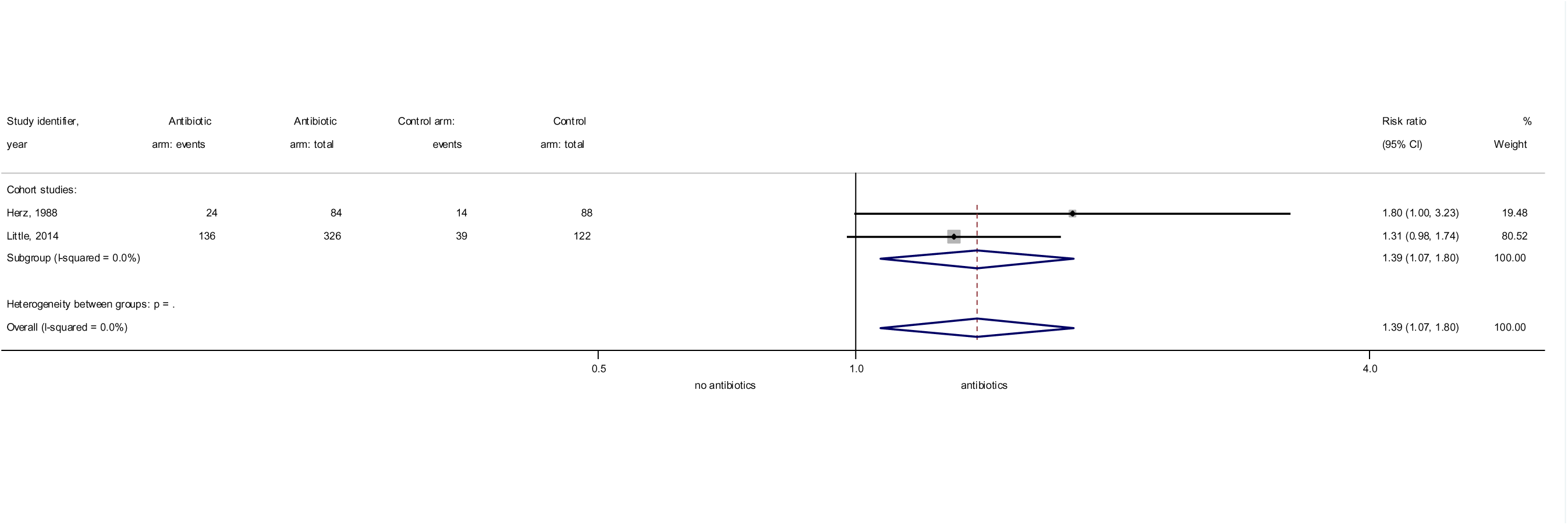
Forest plot of relative risk of reattendance for antibiotics versus no antibiotics, cohort studies excluding Williamson, 2006

## Supplementary tables

**Supplementary Table 1:** Medline search strategy (Ovid MEDLINE® ALL <1946 to 6^th^ Feb 2024>)

| N | Search term | Results |
| --- | --- | --- |
| 1 | (respiratory tract adj3 infection\$).ti,ab. | 27723 |
| 2 | (respiratory tract adj3 infection\$).mp. [mp=title, book title, abstract, original title, name of substance word, subject heading word, floating sub-heading word, keyword heading word, organism supplementary concept word, protocol supplementary concept word, rare disease supplementary concept word, unique identifier, synonyms, population supplementary concept word, anatomy supplementary concept word] | 59888 |
| 3 | exp Pharyngitis/ | 16725 |
| 4 | acute pharyngitis.ti,ab. | 630 |
| 5 | exp Laryngitis/ | 4119 |
| 6 | acute laryngitis.ti,ab. | 198 |
| 7 | sore throat.mp. or Pharyngitis/ | 13662 |
| 8 | sore throat.ti,ab. | 6965 |
| 9 | exp Epiglottitis/ | 1053 |
| 10 | acute epiglottitis.ti,ab. | 617 |
| 11 | exp Tonsillitis/ | 8247 |
| 12 | acute tonsillitis.ti,ab. | 773 |
| 13 | exp Otitis Media/ | 25752 |
| 14 | acute otitis media.ti,ab. | 5436 |
| 15 | exp Otitis Externa/ | 2651 |
| 16 | acute otitis externa.ti,ab. | 158 |
| 17 | exp Sinusitis/ | 23177 |
| 18 | acute sinusitis.ti,ab. | 1202 |
| 19 | exp Rhinitis/ or exp Sinusitis/ | 53209 |
| 20 | acute rhinosinusitis.ti,ab. | 672 |
| 21 | exp Common Cold/ | 4427 |
| 22 | common cold.ti,ab. | 4229 |
| 23 | exp Tracheitis/ | 1560 |
| 24 | acute tracheitis.ti,ab. | 13 |
| 25 | exp Bronchitis/ | 31453 |
| 26 | acute bronchitis.ti,ab. | 1532 |
| 27 | exp Bronchiolitis/ | 9835 |
| 28 | bronchiolitis.ti,ab. | 12586 |
| 29 | 1 or 2 or 3 or 4 or 5 or 6 or 7 or 8 or 9 or 10 or 11 or 12 or 13 or 14 or 15 or 16 or 17 or 18 or 19 or 20 or 21 or 22 or 23 or 24 or 25 or 26 or 27 or 28 | 200078 |
| 30 | exp Anti-Bacterial Agents/ | 814589 |
| 31 | antibiotic*.ti,ab. | 407211 |
| 32 | 30 or 31 | 999639 |
| 33 | reattendance.mp. | 153 |
| 34 | reattendance.ti,ab. | 152 |
| 35 | reconsultation.ti,ab. | 82 |
| 36 | reconsultation.mp. | 85 |
| 37 | further consultation.ti,ab. | 169 |
| 38 | further consultation.mp. | 169 |
| 39 | revisit\$.mp. | 44241 |
| 40 | revisit\$.ti,ab. | 44179 |
| 41 | repeat consult\$.mp. | 43 |

| **N** | **Search term** | **Results** |
| --- | --- | --- |
| 42 | repeat consult\$.ti,ab. | 43 |
| 43 | repeat visit\$.mp. | 405 |
| 44 | repeat attend\$.mp. | 65 |
| 45 | repeat attend\$.ti,ab. | 64 |
| 46 | reconsult\$.mp. | 156 |
| 47 | reconsult\$.ti,ab. | 143 |
| 48 | reattend\$.mp. | 281 |
| 49 | reattend\$.ti,ab. | 280 |
| 50 | consultation rate.mp. | 461 |
| 51 | consultation rate.ti,ab. | 461 |
| 52 | consultation frequency.mp. | 116 |
| 53 | standardised consultation ratio.mp. | 1 |
| 54 | visit rate.mp. | 524 |
| 55 | visit frequency.mp. | 586 |
| 56 | attendance rate.mp. | 1142 |
| 57 | attendance frequency.mp. | 63 |
| 58 | standardised attendance ratio.mp. | 0 |
| 59 | return rates.mp. | 741 |
| 60 | future attendance.mp. | 21 |
| 61 | 33 or 34 or 35 or 36 or 37 or 38 or 39 or 40 or 41 or 42 or 43 or 44 or 45 or 46 or 47 or 48 or 49 or 50 or 51 or 52 or 53 or 54 or 55 or 56 or 57 or 58 or 59 or 60 | 48907 |
| 62 | 29 and 32 and 61 | 147 |
| 63 | Future Likelihood of Seeking care.ti,ab. | 1 |
| 64 | Future Likelihood of Seeking care.mp. | 1 |
| 65 | acute respiratory illness.mp. | 1637 |
| 66 | acute respiratory illness.ti,ab. | 1593 |
| 67 | 65 or 66 | 1637 |
| 68 | 29 or 67 | 201196 |
| 69 | 61 or 63 | 48908 |
| 70 | 32 and 68 and 69 | 148 |

**Supplementary Table 2:**
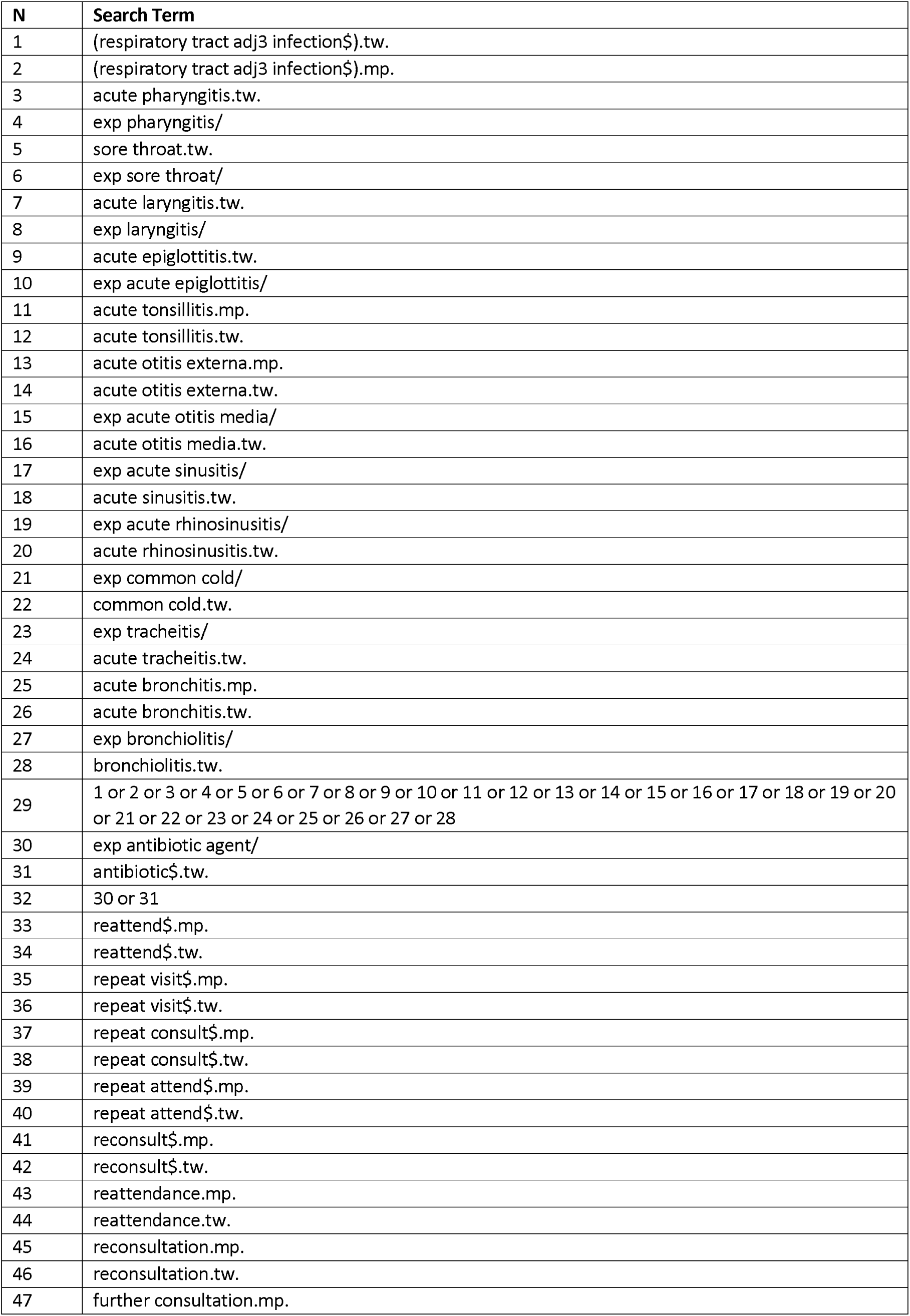
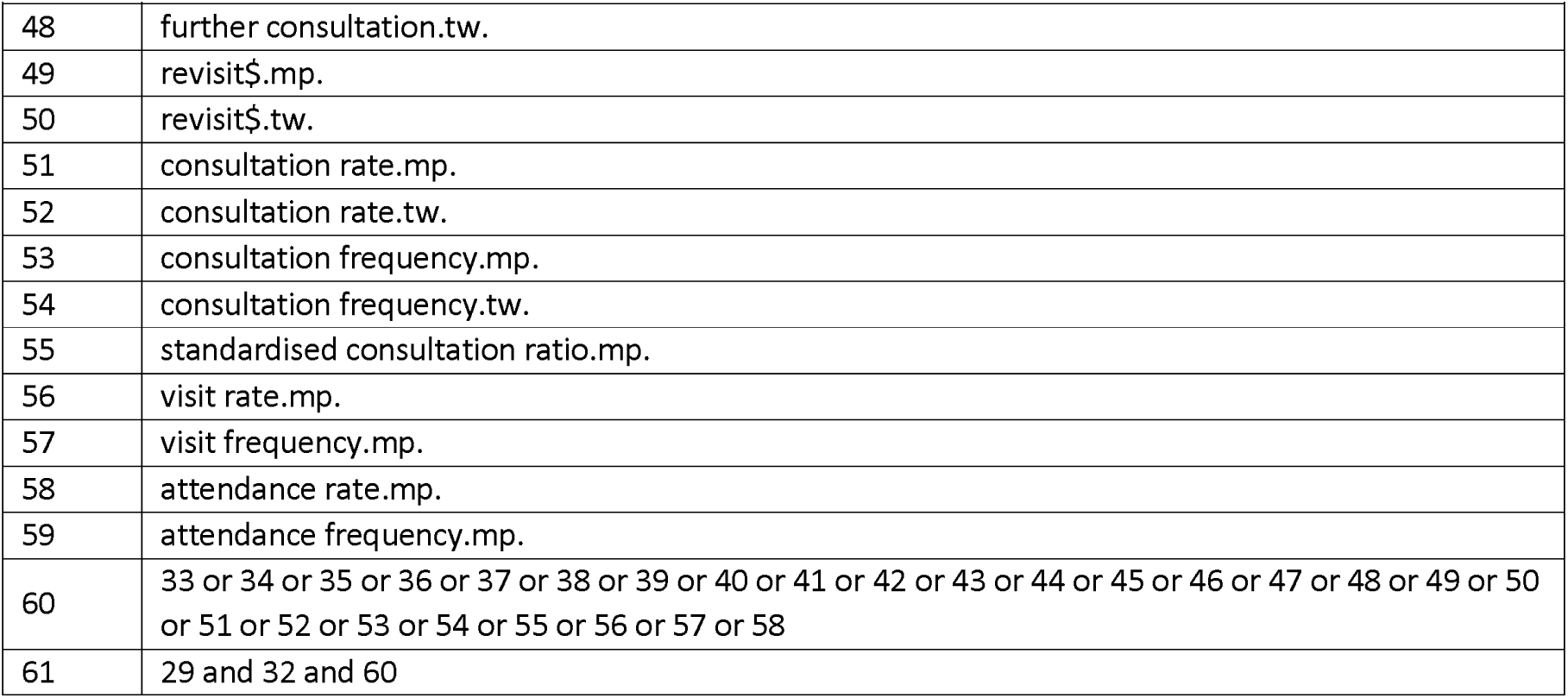
Embase search strategy (Embase <1974 to 6^th^ Feb 2024>)

**Supplementary Table 3:** Search strategy for Cochrane Central.

|  |
| --- |
| COCHRANE CENTRAL 4 MAY 23 2023 |
| respiratory tract adj3 infection\$ OR acute pharyngitis OR sore throat OR acute epiglottitis OR laryngitis OR acute epiglottitis OR acute tonsillitis OR acute otitis media OR acute rhinitis OR acute rhinosinusitis OR common cold OR acute tracheitis OR acute bronchitis OR bronchiolitis AND antibiotic\$ OR reattendance OR reconsultation AND primary care |

**Supplementary Table 4:** Search strategy for Pubmed (MEDLINE)

|  |
| --- |
| Pubmed 4<sup>th</sup> May 2023 |
| ((((((((((((((((((((((respiratory tract adj3 infection\$) ) OR (acute pharyngitis)) OR (sore throat)) OR (acute epiglottitis)) OR (laryngitis)) OR (acute epiglottitis)) OR (acute tonsillitis)) OR (acute otitis media)) OR (acute rhinitis)) OR (acute rhinosinusitis)) OR (common cold)) OR (acute tracheitis)) OR (acute bronchitis)) OR (bronchiolitis)) AND (antibiotic\$) OR (reattendance)) OR (reconsultation)) AND (primary care) |
| Filters: Observational Study, Randomized Controlled Trial |

**Supplementary Table 5:**
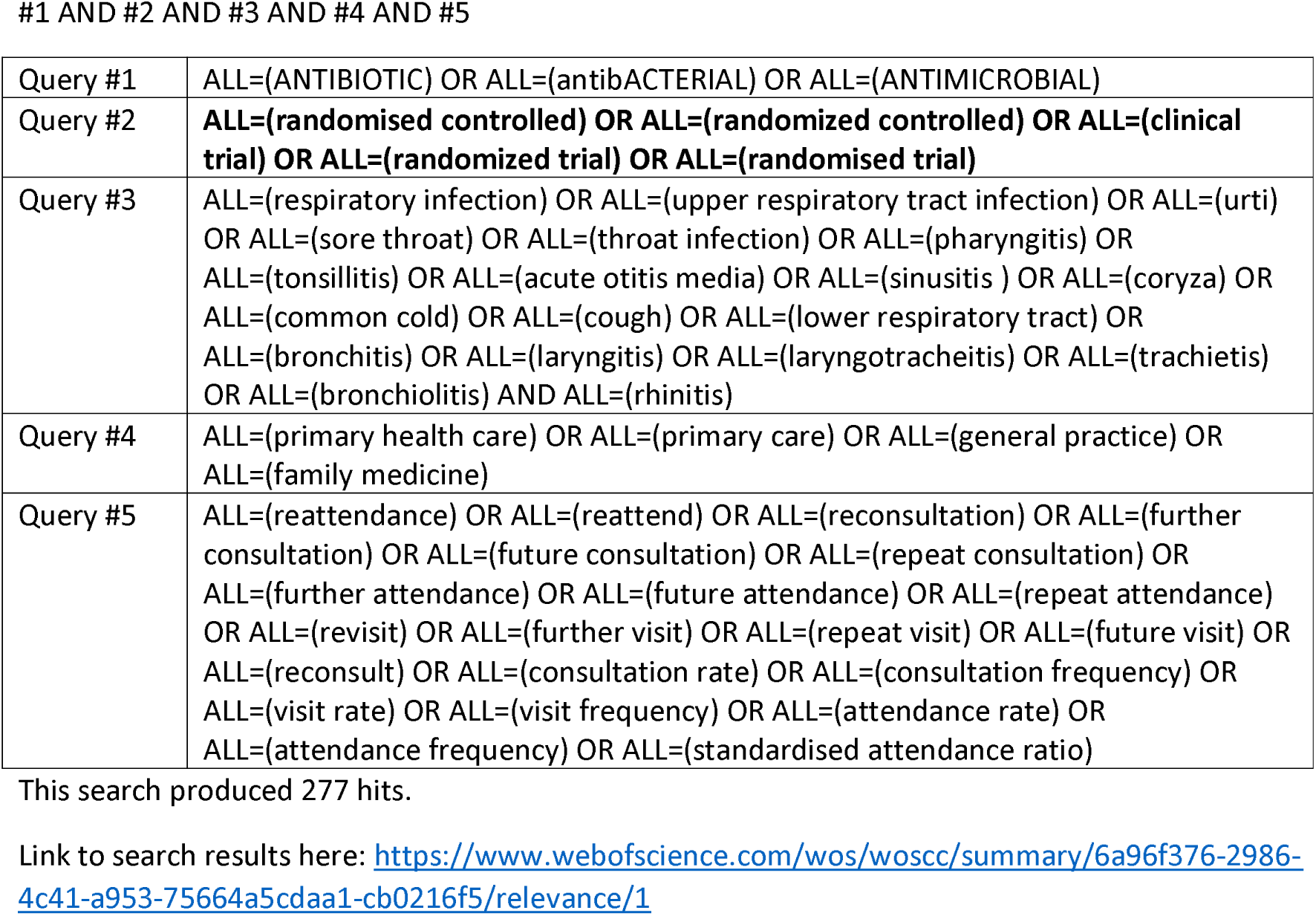
Search in Web of Science Science Citation Index Expanded – 1900 to present Conference proceedings citation index – Science – 1990 to present #1 AND #2 AND #3 AND #4 AND #5.

|  |  |
| --- | --- |
| Query #1 | ALL=(ANTIBIOTIC) OR ALL=(antibACTERIAL) OR ALL=(ANTIMICROBIAL) |
| Query #2 | **ALL=(randomised controlled) OR ALL=(randomized controlled) OR ALL=(clinical trial) OR ALL=(randomized trial) OR ALL=(randomised trial)** |
| Query #3 | ALL=(respiratory infection) OR ALL=(upper respiratory tract infection) OR ALL=(urti) OR ALL=(sore throat) OR ALL=(throat infection) OR ALL=(pharyngitis) OR ALL=(tonsillitis) OR ALL=(acute otitis media) OR ALL=(sinusitis ) OR ALL=(coryza) OR ALL=(common cold) OR ALL=(cough) OR ALL=(lower respiratory tract) OR ALL=(bronchitis) OR ALL=(laryngitis) OR ALL=(laryngotracheitis) OR ALL=(trachietis) OR ALL=(bronchiolitis) AND ALL=(rhinitis) |
| Query #4 | ALL=(primary health care) OR ALL=(primary care) OR ALL=(general practice) OR ALL=(family medicine) |
| Query #5 | ALL=(reattendance) OR ALL=(reattend) OR ALL=(reconsultation) OR ALL=(further consultation) OR ALL=(future consultation) OR ALL=(repeat consultation) OR ALL=(further attendance) OR ALL=(future attendance) OR ALL=(repeat attendance) OR ALL=(revisit) OR ALL=(further visit) OR ALL=(repeat visit) OR ALL=(future visit) OR ALL=(reconsult) OR ALL=(consultation rate) OR ALL=(consultation frequency) OR ALL=(visit rate) OR ALL=(visit frequency) OR ALL=(attendance rate) OR ALL=(attendance frequency) OR ALL=(standardised attendance ratio) |

This search produced 277 hits.
Link to search results here: https://www.webofscience.com/wos/woscc/summary/6a96f376-2986-4c41-a953-75664a5cdaa1-cb0216f5/relevance/1

**Supplementary Table 6:** General information & selection criteria.

|  |  |  |
| --- | --- | --- |
| Study title |  |  |
| Study ID (surname of first author + year of publication) |  |  |
| ID of person extracting data (Initials) |  |  |
| Study author's contact details |  |  |
| Publication type (full report or abstract) |  |  |
| Date form was completed (dd/mm/yyyy) |  |  |
| Study characteristics | Eligibility criteria | Eligibility criteria met.<br>(Yes) (No) (Unclear) |
| Study design | Randomised controlled trial (unblinded)<br>Cohort study |  |
| Population | Adults or children with respiratory tract infections presenting in primary care |  |
| Intervention (exposure) | Antibiotics (immediate or delayed) |  |
| Comparison | No antibiotics |  |
| Outcome | Future reattendance: ≥28 days / 1 month |  |
| Setting | Primary care |  |
| Include<br>(Tick as appropriate) |  | Exclude<br>(Tick as appropriate) |
| Reasons for exclusion if applicable |  |  |

**Supplementary Table 7:** Characteristics of included studies.

|  | Description as stated in the report or paper | Location in the text |
| --- | --- | --- |
| Aim of the study |  |  |
| Study design |  |  |
| Start date |  |  |
| End date |  |  |
| Study duration |  |  |
| Sample size |  |  |
| Country of origin |  |  |
| Ethical approval needed/obtained for the study |  |  |
| Characteristics of the population |  |  |
| Inclusion criteria |  |  |
| Exclusion criteria |  |  |
| Type of intervention (immediate or delayed antibiotic prescription) |  |  |
| Information on the outcome (estimate measure, unit of measurement, dichotomous or continuous variable) |  |  |
| Source of funding |  |  |
| Conflict of interest |  |  |

